# Relative humidity predicts day-to-day variations in COVID-19 cases in the city of Buenos Aires

**DOI:** 10.1101/2021.01.29.21250789

**Authors:** Andrea L. Pineda Rojas, Sandra M. Cordo, Ramiro I. Saurral, Jose L. Jimenez, Linsey C. Marr, Emilio Kropff

**Affiliations:** CIMA, UMI-IFAECI/CNRS, FCEyN, Universidad de Buenos Aires – UBA/CONICET, Buenos Aires, Argentina; Laboratorio de Virología, QB, FCEyN, Universidad de Buenos Aires – IQUIBICEN/CONICET, Buenos Aires, Argentina; DCAO, FCEyN, Universidad de Buenos Aires, Buenos Aires, Argentina; Dept. of Chemistry and CIRES, University of Colorado, Boulder, CO, USA; Civil and Environmental Engineering, Virginia Tech, Blacksburg, Virginia, USA; Leloir Institute – IIBBA/CONICET, Buenos Aires, Argentina

**Author notes:** Corresponding Author Emilio Kropff, Leloir Institute – IIBBA/CONICET, Av. Patricias Argentinas 435, CBA, Buenos Aires.

**Keywords:** COVID-19, Relative humidity, airborne transmission, meteorology

## Abstract

Possible links between the transmission of COVID-19 and meteorology have been investigated by comparing positive cases across geographical regions or seasons. Little is known, however, about the degree to which environmental conditions modulate the daily dynamics of COVID-19 spread at a given location. One reason for this is that individual waves of the disease are typically too abrupt, making it hard to isolate the contribution of meteorological cycles. To overcome this shortage, we here present a case study of the first wave of the outbreak in the city of Buenos Aires, which had a slow evolution of the case load extending along most of 2020. We found that humidity plays a prominent role in modulating the variation of COVID-19 positive cases through a negative-slope linear relationship, with an optimal lag of 9 days between the meteorological observation and the positive case report. This relationship is specific to winter months, when relative humidity predicts up to half of the variance in positive cases. Our results provide a tool to anticipate local surges in COVID-19 cases after events of low humidity. More generally, they add to accumulating evidence pointing to dry air as a facilitator of COVID-19 transmission.

## INTRODUCTION

Increasing evidence points to aerosols as a main mode of transmission of COVID-19, mostly occurring indoors^1-3^. Studies suggest that environmental conditions may affect airborne transmission of respiratory viruses through different processes^4^. First, temperature (T) and relative humidity (RH) can modulate through evaporation the size distribution of exhaled aerosol particles, determining the number of particles that does not settle due to gravity and can stay suspended in the air^5, 6^. Second, they can also influence the buoyancy of the exhaled respiratory plume (which is a mixture of gases and aerosols) determining if and how it rises^7^. Third, T and RH affect the decay rate of the virus inside aerosols and droplets^6, 8^. Fourth, humidity is known to affect the immune response of the respiratory system^9^. Fifth, ambient conditions including air flow and turbulence affect transport and dispersion of the respiratory plume, and therefore the aerosol concentration at a given distance from an infected person^10^. In addition, environmental conditions may influence human behavior, such as the amount of time spent outdoors or ventilation patterns affecting the accumulation of aerosols indoors.

Massive monitoring of indoor air conditions would be ideal to understand which of these variables affects COVID-19 transmission at the population level. In the absence of this kind of data, however, widely available outdoor meteorology can be used as a proxy of indoor conditions, albeit mediated by levels of heating and ventilation^6^. Since the start of the pandemic, several studies have assessed the relationship between the number of daily confirmed COVID-19 cases (positive cases) and meteorological conditions in different regions of the world ^11-17^. Variables such as humidity, temperature, solar radiation, precipitation or wind speed have been found to co-vary with positive cases. Two approaches have been typically followed. The first is to compare the spread of the disease across geographical regions. In this kind of study, variability in meteorological conditions is achieved by including enough spatially distant locations, but differences other than meteorological between locations introduce heterogeneity. The second approach is to correlate the initial growth phase of the outbreak on a single location with a number of meteorological variables. The challenge in this kind of study relates to the fact that many meteorological variables have marked seasonal variations, and correlations with an abrupt increase in the number of positive cases might be spurious, simply reflecting two parallel but independent trends. In addition, the dynamic of local mitigation policies and the response of the population represent confounding or heterogeneity factors for both approaches. Due to these shortcomings, the question of whether or not meteorology can modulate COVID-19 spread is open, with sensible arguments on both sides^18, 19^.

Here we present a study of the influence of meteorological variables during the first wave of spread (8-months) of COVID-19 in the city of Buenos Aires (CBA), Argentina. CBA has a humid subtropical climate with four distinct seasons, and is a useful case study for several reasons. First, over 5% of its 2.9 million inhabitants have suffered from COVID-19 in 2020, as confirmed by PCR tests, during a single long wave of the spread starting at the beginning of May and reaching its peak by the end of August (Figure 1A). Second, although the area of the city is considerable (209 km^2^), it is located on a flat terrain, so that surface meteorological conditions can be assumed to be horizontally homogeneous^20^. Third, in 2020 the most disruptive policies were enforced when there were still fewer than 50 daily cases (lockdown: March 20^th^; facemasks mandatory in public spaces including shops, etc.: April 15^th^, 2020) in the hope of flattening the curve. These measures were renewed roughly every 15 days, effectively extending the lockdown until November 9^th^, 2020, a period that included the whole austral winter (June to September).

**Figure 1.**
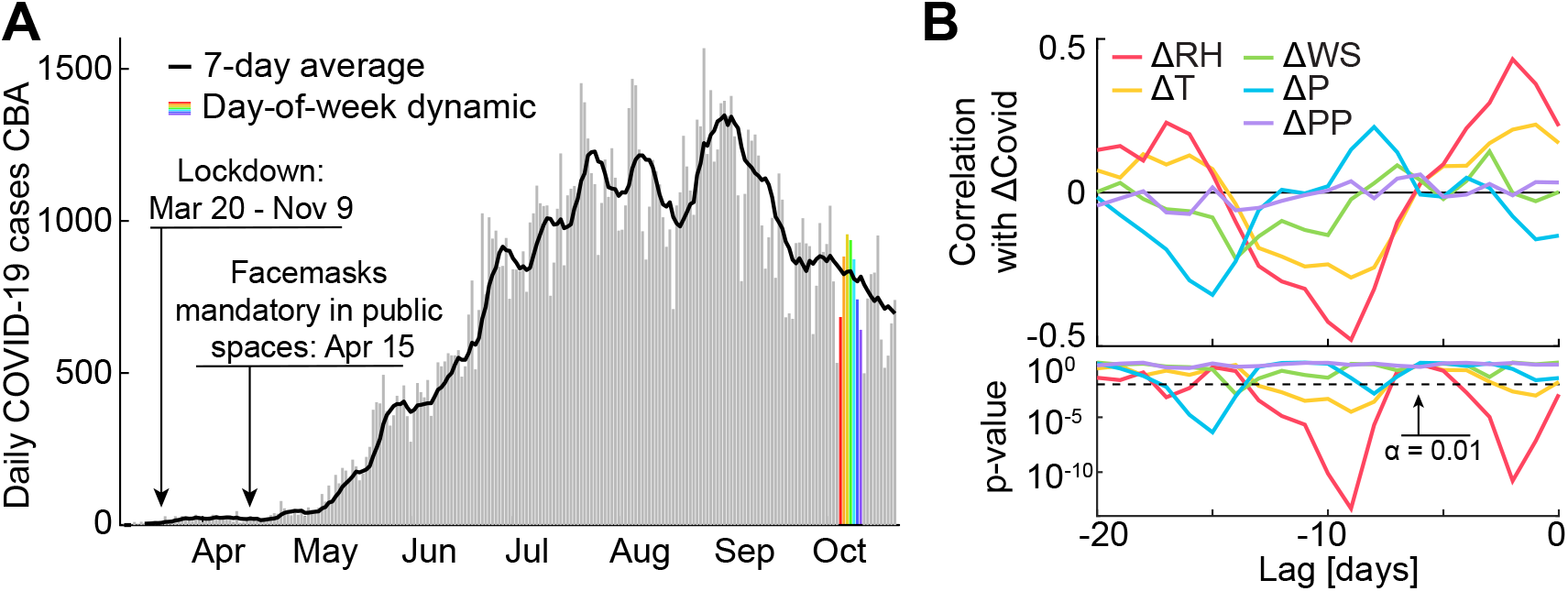
(A) Daily cases in CBA taken from dataset #1 (grey bars), 7-day average (black) and an example of the fast day-of-week dynamic (colors). Enforcement of major policy changes indicated. (B) Top: Pearson correlation between *ΔCovid(t)* and meteorological counterparts (color coded), for different lags expressed in days. Bottom: Corresponding p-values (log scale; same color code).

Our work aims to identify meteorological variables that modulate day-to-day variations of the COVID-19 spread in CBA. We introduce a simple conceptualization that allows filtering out the main driver of the spread to focus on such modulations. To disentangle the effect of highly inter-correlated meteorological variables we use cross-validation techniques. Finally, we show how these simple tools can be used to predict variations in the number of COVID-19 cases, which could be useful for health institutions to plan logistics around a week in advance.

## MATERIALS AND METHODS

### Data gathering

Two datasets of confirmed COVID-19 cases (SARS-CoV2 positive PCR tests) in CBA during the period March to November, 2020 were used (Figure S1):

i. dataset #1 includes the date of report of all known positive cases, from the daily reports by Health Ministry of Argentina, compiled and curated by Sistemas Mapache (github.com/SistemasMapache/Covid19arData), and
ii. dataset #2 has the date in which symptoms began for only 60% of positive cases, from the Health Ministry of Argentina open data webpage (datos.salud.gob.ar/dataset)

Most of the study was performed for the complete dataset #1, while dataset #2 was used to complement the analysis. All data were processed through custom scripts written in MATLAB (MathWorks, Natick, MA).

Meteorological data corresponding to the station located at the domestic airport (AEP; World Meteorological Organization station number 87582) for the same period was obtained from public databases provided by the National Oceanic and Atmospheric Administration (NOAA, US)^21^ and OGIMET (www.ogimet.com). We included as meteorological variables for our study the surface daily mean values of relative humidity (RH), temperature (T), wind speed (WS), pressure (P), precipitation (PP) and sky cover (SC), as well as the surface daily minimum (T_min_) and maximum (T_max_) values of hourly temperature. In addition, absolute humidity (AH) was estimated from RH and T using the formula^22^:

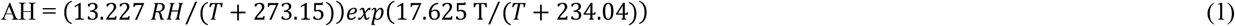

### Isolating the mid-range dynamic

The curve of daily new COVID-19 cases in CBA exhibits different scales of temporal variability (Figure 1A, dataset #1). As in many such datasets, fast and slow dynamics are observed. The slow dynamic reflects the main waves of spread of the pandemic, which in the particular case of CBA took during most of 2020 the form of a single, slowly modulated wave starting in May and reaching its peak in August. On the opposite extreme of the spectrum, the fast dynamic reflects day-of-week fluctuations (especially weekend vs. weekday differences). In addition, a third, intermediate dynamic range is apparent in this dataset, represented by fluctuations with an irregular periodicity in the range of 2-4 weeks. In the present work, we characterized these mid-range fluctuations following the hypothesis that they are at least in part rooted on meteorological conditions. This would not imply that meteorological conditions are the main driver of the spread, but rather that they can modulate transmission. To study these mid-range variations, filtering out the slow (main driver) and fast (day-of-week) dynamics, we defined the weekly difference of any given variable *X(t)* as the value taken by this variable on day *t* minus its value 7 days before:

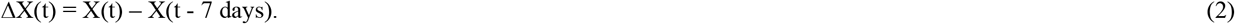

Equation (2) was applied to daily COVID-19 positive cases to obtain *ΔCovid(t)* and to its meteorological counterparts (e.g., *ΔRH(t)* for relative humidity). To study the relationship between these variables we used linear models and the Pearson correlation coefficient, including a range of lags between the day of the meteorological observation and that of the COVID-19 positive case report. Significance for a linear relationship against the null hypothesis of no improvement over a constant model was obtained as a p-value through a t-test using the MATLAB function *fitlim()*. The optimal lag for a given variable was defined as the lag with both minimum p-value and maximum absolute correlation with *ΔCovid(t)*.

Note that the Δ transformation introduced in Equation 2, while convenient for isolating the mid-range dynamic, has a small disadvantage. Since a given value of RH(t) impacts on both ΔRH(t) and ΔRH(t+7), a strong correlation at a given lag is typically mirrored by moderately strong correlations of opposite signs at lags 7 days earlier or later. To avoid any ambiguity, in all cases the optimal lag was determined by the global maximum of absolute correlation, or equivalently (since degrees of freedom do not change) the global minimum of p-value, regardless of other local extrema. Furthermore, while the Δ transformation is central to our results, an independent analysis not utilizing it was applied to the same data to confirm the typical size of optimal lags.

### Testing day-of-week modulation

Data of *Covid(t)* and *ΔCovid(t)* were normalized by the 7-day average of *Covid(t)* and divided into 7 groups, according to the day of the week of *t* (Figure S2). The Kruskal-Wallis test was used to assess potential differences between groups indicative of day-of week modulation.

### Optimization of the multivariate linear model

Cross-validation allows for the fair comparison between models of different complexity^23^. We performed full 10-fold cross-validation including all possible combinations of the 8 meteorological variables included in the study. All linear models with a number of variables ranging from 0 (constant model) to 8 (all variables included) were compared. For each combination of variables, the data was divided into 10 random subgroups of equal size. Each subgroup was once set aside and used as a test set for the linear model trained with the remaining 9 groups. For a given number of variables (0 to 8), the best model was chosen as the one minimizing the test sum of squared residuals. To choose between models with different number of variables, the one-standard-error criterion was used^23^. This criterion defines the best model as the one with fewest variables that has a test sum of squared residuals not higher than the lowest one plus one standard error. All linear combinations of variables and interactions between variables were tested for two variations of the data: a) each variable with its own optimal lag and b) all variables using the optimal lag for *ΔRH*.

To corroborate the results, we repeated all these analyses using the MATLAB *lasso()* function. The Lasso method implements the choice of the optimal model by minimizing squared residuals while penalizing non-zero linear coefficients (i.e., the effective number of terms in the model) under the one-standard-error criterion. This is achieved through a cost function defined as the sum of the absolute value of linear coefficients ^23^.

Cross-validation and Lasso were also used to specifically disentangle the relative contribution of *ΔAH, ΔRH* and *ΔT* in explaining *ΔCovid* variability, given that laboratory experiments suggest that these variables modulate virus survival in aerosols^6^.

### Testing non-linear relationships

Given that meteorological variables have been described to sometimes exhibit a non-linear relationship with positive COVID-19 cases ^24^, we also tested non-linear correlations (Spearman) and non-linear models (up to 4 degree polynomials including all possible interactions between variables). However, Spearman correlations were very similar to Pearson correlations (Figure S3), with identical optimal lags for all variables exhibiting a significant correlation. In addition, the two cross-validation methods that we used indicated that non-linear terms offered no improvement over the linear model. For these reasons, our work mostly presents the results obtained within the linear framework.

## RESULTS AND DISCUSSION

### Meteorology anticipates changes in COVID-19 transmission

The time series of COVID-19 confirmed cases in dataset #1 exhibited, in addition to the main driver and the day-of-week modulations, a mid-range dynamic of variability, with an unstable period of around 2 to 4 weeks (Fig. 1A). To understand if meteorological factors explained this mid-range modulation, we applied Equation 2 to all variables, obtaining *ΔCovid(t)* and its meteorological counterparts (e.g., *ΔRH(t)* for relative humidity). As expected, there was no significant day-of-week modulation in *ΔCovid(t)* (Figure S2; Kruskal-Wallis test for H_0_ defined by no difference between days of the week: χ_2_(6) = 0.42, p = 0.99). We studied the Pearson correlation between *ΔCovid(t)* and its meteorological counterparts (Figure 1B; see also Figures S3 and S4). Since meteorology is not expected to affect the outcome of positive COVID-19 cases reported on the same day, but rather those reported some days later, lags ranging from 0 to 20 days were included in the analysis. We found that several variables significantly correlated with *ΔCovid(t)* at different lags, the most frequent of which was 9 days (Figure S3). It should be noted that in dataset #1 these lags aim to capture an average time window corresponding to the incubation period plus time for testing and bureaucratic processing of data. Correlations and lags were similar when the Spearman coefficient was used, suggesting that the linear framework, while simpler, correctly captures the individual relationships between *ΔCovid(t)* and meteorological counterparts. Of all variables, *ΔRH(t – 9 days)* exhibited the most extreme correlation with *ΔCovid(t)* (Pearson correlation: -0.48; t(218): -8.1; p: 10^−13^). The evolution of *ΔCovid(t)* was strikingly similar to that of negative *ΔRH(t – 9 days)* (Figure 2), especially so when considering the numerous confounders or sources of heterogeneity expected to obscure a relationship between meteorology and the reporting of COVID-19 positive cases (individual variability in symptomatic response, test processing time or RH indoor-outdoor relationship to cite only a few).

**Figure 2.**
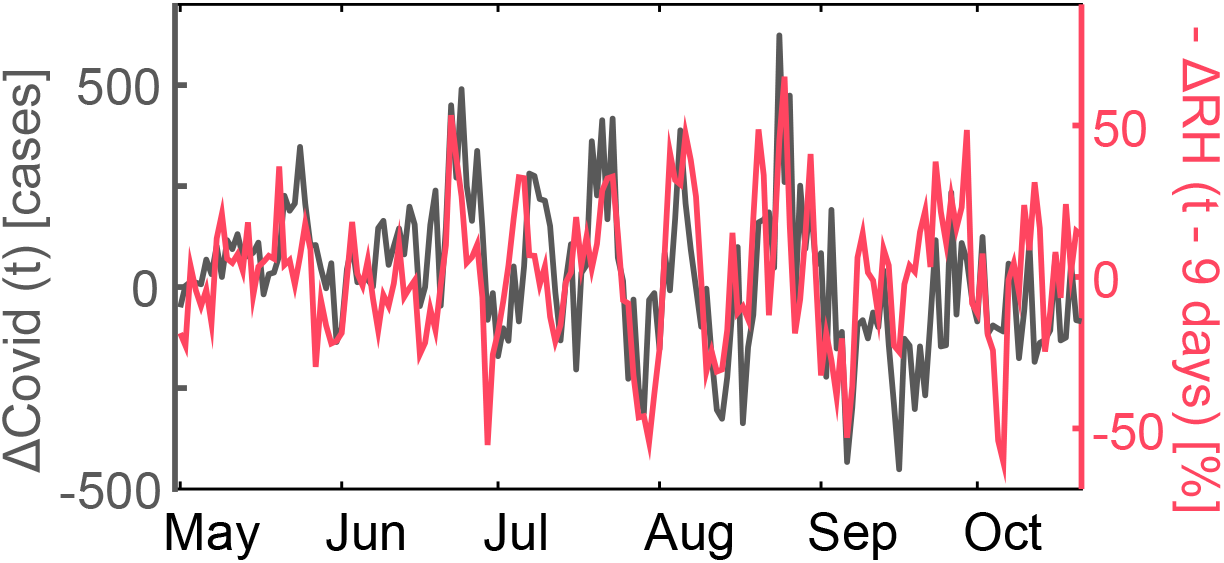
Co-evolution of *ΔCovid(t)* (grey; left axis) and negative *ΔRH(t – 9 days)* (red; right axis).

### Variables other than RH play a negligible role

Meteorological variables are tightly interrelated. To assess the relative importance of *ΔRH* compared to the other potential predictors of *ΔCovid*, we applied two cross-validation methods (full 10-fold cross-validation and Lasso). These methods yield as an output the linear combination of predictors that best model *ΔCovid(t)*, avoiding contributions that are negligible or related to overfitting. Both methods led to an identical conclusion, pointing to

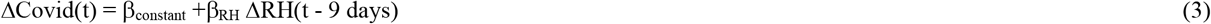

as the best model of *ΔCovid(t)*, where *β_X_* stands for the linear coefficient for variable *X*.

While evidence from laboratory experiments suggests an important role played by RH in modulating the efficacy of transmission of airborne viruses, AH has been pointed to be in some situations better than RH as a proxy for indoor RH^6^. The reason for this is that indoor and outdoor AH are in general similar, but potential differences between indoor and outdoor T due to heating can have a large impact on indoor RH (see Equation 1 for an approximate relationship between these variables). This might depend on the type of construction and heating systems prevalent at a given location, and has not been studied specifically in CBA. Although we did not have an independent measure of AH, we used Equation 1 to estimate it, and repeated the optimal lag search (Figure S5) and the cross-validation procedures using exclusively *ΔRH, ΔT* and *ΔAH*. Again, both methods showed that Equation 3 represents the best model of *ΔCovid*.

### RH anticipates variations in the triggering of symptoms

In dataset #1, the optimal lag of 9 days includes the incubation period plus time for testing and bureaucratic processing of data. To dissect this time window, we analyzed dataset #2 in a similar way. This dataset included, for around 60% of confirmed positive cases, the date in which symptoms began. Defining *CovidS(t)* as the number of confirmed cases with symptoms triggered at a given day (Figure S1), we obtained *ΔCovidS(t)* through Equation 2 and found that its optimal lag relative to *ΔRH* was of 5 days (Pearson correlation: -0.44; t(218): -7.3; p: 5 × 10^−12^). Since 5 days is the average window for the development of COVID-19 symptoms^25^, this further supports a specific role played by humidity in early stages of the disease. The rest of this work uses dataset #1, which includes all confirmed cases.

### RH modulates transmission only during winter

Given that in CBA a single wave of the pandemic extended across seasons, we next assessed whether or not a seasonal effect modulated the *ΔCovid* -*ΔRH* relationship. We classified the data by month of the COVID-19 report date (*t*) from March to October, 2020. For each month, we plotted *ΔCovid(t)* vs. *ΔRH(t – 9 days)* and obtained the individual slopes (linear coefficient *β_RH_* in Equation 3) and correlation coefficients (Figure 3). We observed that the modulation of *ΔCovid* by *ΔRH* was only highly significant during the winter months (June to August), with similar values of *β_RH_* (Figure 3A). For transition months (May and September) the relationship weakened toward not significant, regardless of whether the overall number of COVID-19 positive cases was low (as in April) or high (as in October). Given that cross-validation procedures showed no significant interactions, this seasonal effect cannot be explained by the meteorological variables considered here. Mean monthly temperature, however, correlated with *β_RH_* across months (Pearson correlation: 0.88; t(6): 4.6; p: 4 × 10^−3^). We speculate that rather than causality, this correlation could reflect indirect mechanisms such as seasonal changes in people behavior (e.g., habits regarding ventilation, the use of heating or outdoor vs. indoor gathering).

**Figure 3.**
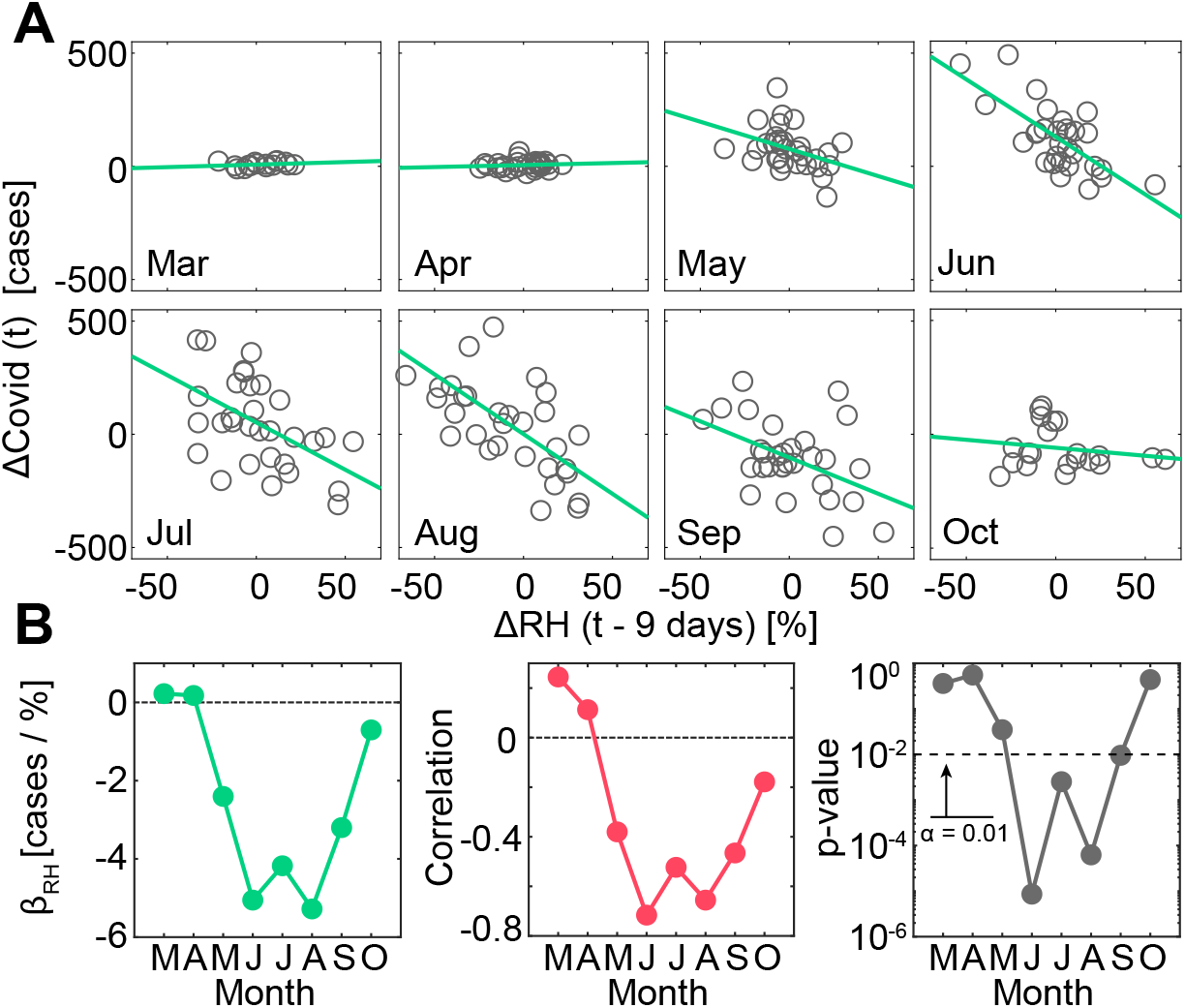
(A) Plot of *ΔCovid(t)* vs *ΔRH(t – 9 days)* (grey; one circle per day) across individual months (one month per sub-panel) together with linear fits (green). (B) Slope (left), Pearson correlation coefficient (center) and corresponding p-value (right) across months, obtained from plots in (A).

### RH as a tool to anticipate an increase in positive cases

Our results could provide a tool to anticipate local surges in COVID-19 due to low *RH*. To demonstrate this, we first studied the average evolution of *RH* in anticipation of events of an extreme increase in the number of positive cases (Figure 4A). We identified 7 peaks in *ΔCovid* that were higher than 200 positive cases and studied the evolution of RH during the 20 days prior to them. A trough in *RH* extending roughly from days 15 to 5 prior to the peak in *ΔCovid*, and reaching an average value of 55% at its minimum, was observed. This confirms that a low value of humidity precedes an increase in the number of positive cases with a lag similar to the one found using Pearson correlations.

**Figure 4.**
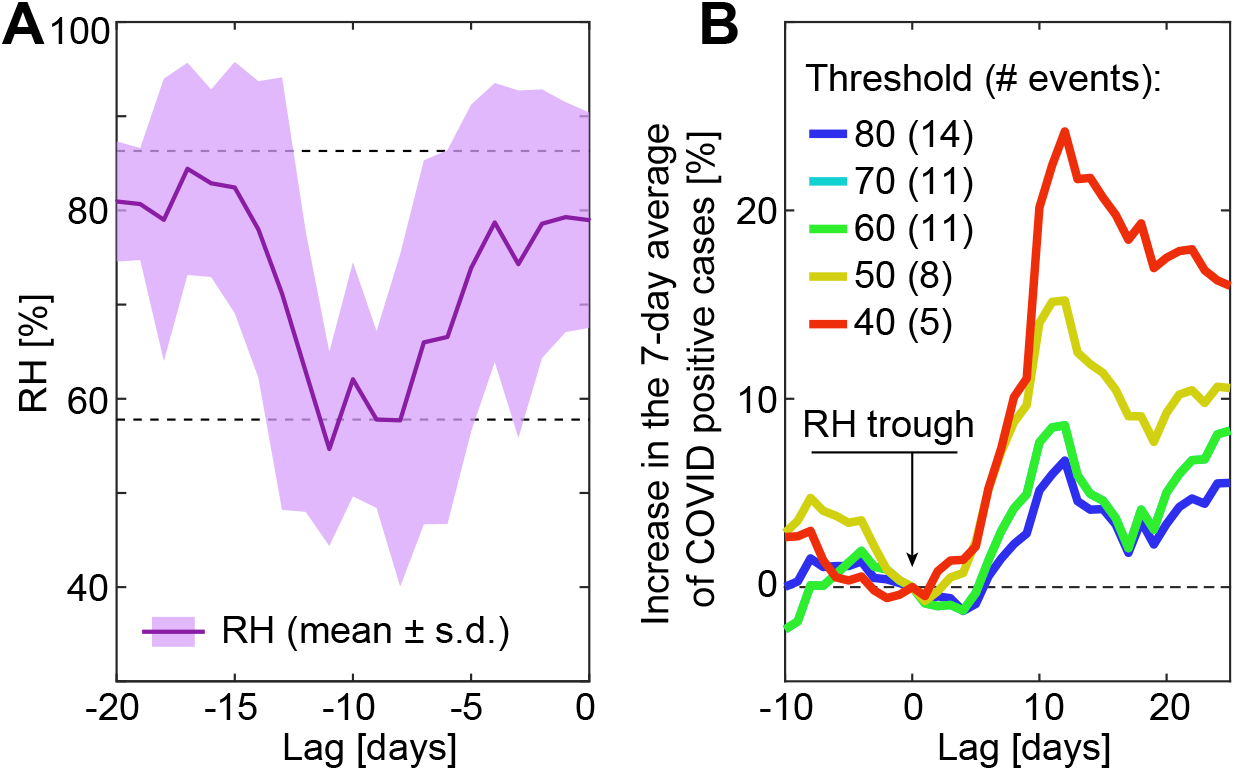
(A) Distribution (mean ± s.d.) of RH during the 20 days prior to peaks of *ΔCovid* higher than 200 cases (7 events). Dashed lines: mean ± s.d. of RH for the whole dataset (March to October, 2020). (B) Mean evolution of the 7-day average of COVID-19 cases relative to the value at minima of RH (lower than color coded threshold; number of events indicated).

Inversely, we asked what was the evolution of COVID-19 positive cases around extreme low values of RH (Figure 4B). For a given humidity threshold (a range going from 40% to 80% was studied), we identified the throughs in RH that went below threshold. We then studied the percentage of change in the 7-day average of COVID-19 positive cases relative to the value observed on the day of the RH through (zero lag). We observed an average increase in COVID-19 cases starting 5 to 7 days after troughs of RH of different magnitude and peaking around day 11. For the 5 lowest humidity events (< 40%) that took place during the period under study, this implied an average increase of more than 20% in reported COVID-19 cases. This result corroborates our main findings without making use of the transformation of the data introduced in Equation 2, suggesting that widely available meteorological observations of RH can be informative to anticipate surges in the number of COVID-19 cases.

## CONCLUSIONS

This study provides a fresh perspective on the day-to-day dynamic of the COVID-19 pandemic. The methodology that we introduce for the first time in this kind of study (Equation 2) allowed us to inspect, with minimal intervention over the raw data, mid-range modulations in the dynamic of positive COVID-19 cases, filtering out at the same time the main driver of the spread and the day-of-week effect. Several variables that are critical to understand the dynamics of the main driver, such as testing capacity, positivity or response of the population to guidelines, were not considered here. However, they are unlikely to significantly affect our results since they reflect slow variations that are filtered out by Equation 2. In addition, the only situation in which these variables could explain the observed correlation between *ΔCovid* and *ΔRH* is if they were themselves related to humidity, which is highly unlikely.

Our main result is that changes in daily RH anticipate changes of opposite sign in the number of COVID-19 positive cases observed 9 days later in CBA. An analysis of a subset of the data indicates that this 9-day window can be divided into a 5-day incubation period plus a 4-day testing and data processing period. Although other meteorological variables exhibited similar (weaker) relationships, our cross-validation procedures indicated that this was due to the complex set of interactions between meteorological variables. RH alone was as good in describing variations in positive cases as the whole set of variables. When considering potential seasonal effects, we observed that the linear relationship between variations of RH and positive COVID-19 cases was only significant during winter months. During this period, sustained monthly levels of correlation and slope were found, smoothly varying toward zero slope in the transition months. This rules out anecdotal correlations, and rather makes it likely that some behavioral pattern consistently occurring during winter, for example reduced levels of ventilation or the use of heating, is necessary for the modulation of COVID-19 transmission by RH. Our findings provide a practical tool to predict a raise of up to 20% in positive cases following extreme low values of RH during winter months, which could be useful for the planification of logistics in health institutions of CBA.

Further efforts should be directed to understand which of our results can be replicated in other locations of the world and which are specific to the CBA outbreak (and why). In addition, information regarding the type of transmission in each case (close proximity vs shared-room scale), if available, could shed light on the mechanisms behind our results. Evaporation and buoyancy act within the first seconds of release of exhaled aerosols, while viral decay typically requires longer timescales^26^. Hence, a modulation of COVID-19 transmission by RH found exclusively in cases of shared-room scale would indicate that the main mechanism behind it is viral decay. In contrast, if other mechanisms contributed significantly to the effect, both types of transmission would exhibit similar levels of modulation. Given that airborne transmission mechanisms are shared by different respiratory viruses, developments in this area would improve our understanding not only of the current pandemic, but possibly of other diseases that pose a threat to human health and welfare.

## Data Availability

This manuscript is based on public data. Sources are listed in Materials and Methods.

## AUTHOR INFORMATION

### Author Contributions

The manuscript was written through contributions of all authors. / All authors have given approval to the final version of the manuscript.

### Funding Sources

#### Notes

Any additional relevant notes should be placed here.

## ACKNOWLEDGMENT

The work was supported by PICT 2015-1273 (E.K.), PICT 2018-1624 (A.P.R.) and PICT 2017-1136 (S.C.) grants from the Science Ministry of Argentina.

## ABBREVIATIONS

CBA: city of Buenos Aires
RH: relative humidity
AH: absolute humidity
T: temperature
WS: wind speed
P: pressure
PP: precipitation
SC: sky cover.

---

**Figure S1.**
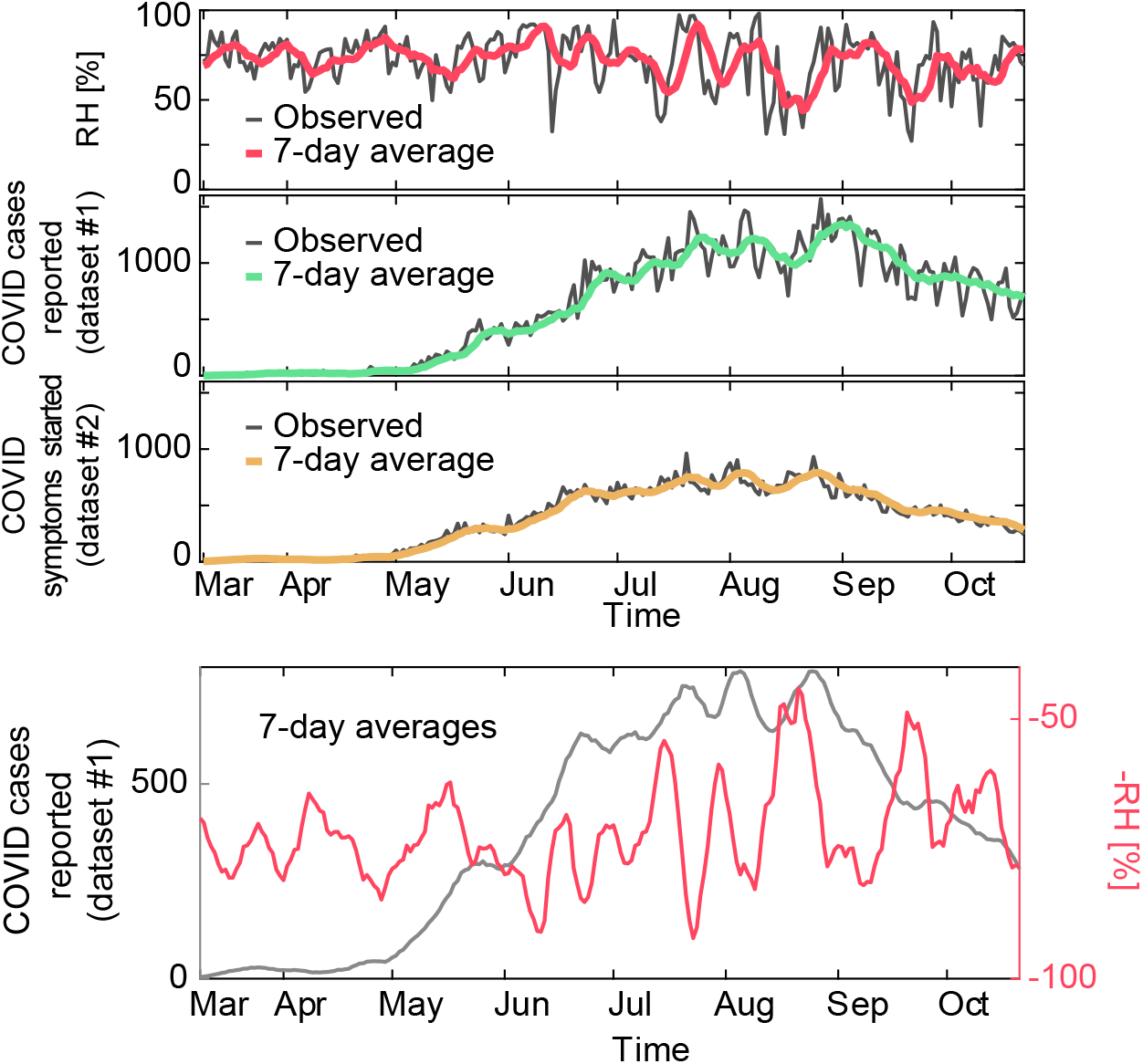
Top panel: Time series (grey) and 7-day average (color) of RH (top) Covid(t) (middle; dataset #1) and CovidS(t) (bottom; dataset #2). Bottom panel: 7-day average of Covid(t) (left axis, grey) and negative RH (right axis, red).

**Figure S2.**
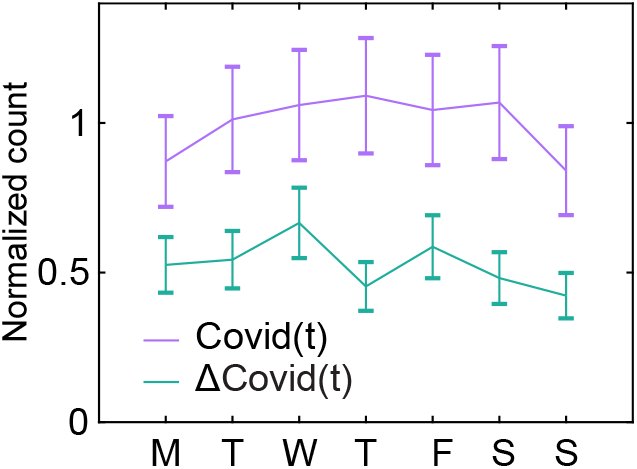
Distribution (mean ± s.e.m.) of *Covid(t)* (purple) and *ΔCovid(t)* (green) according to the day of the week of t. Both series were normalized by the 7-day average of *Covid(t)* to account for the variability across months, which otherwise masked the day of week variability. The Kruskal Wallis test showed a significant day-of-week effect only for *Covid(t)* (*Covid*: χ^2^(6) = 43.9, p = 10^−7^; *ΔCovid*: χ^2^(6) = 0.42, p = 0.99)

**Figure S3.**
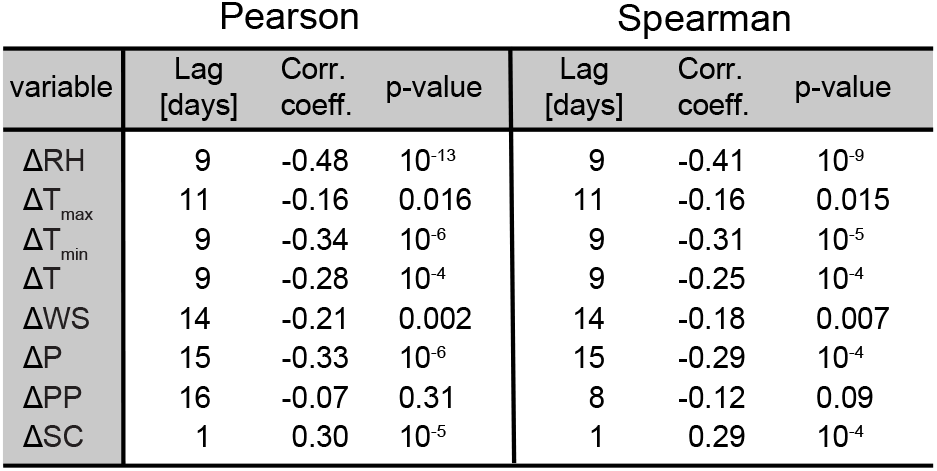
For all meteorological variables, optimal lag and corresponding correlation using either the Pearson or Spearman coefficients. For each type of correlation columns show, from left to right, optimal lag expressed in days, correlation coefficient at the optimal lag and associated p-value.

**Figure S4.**
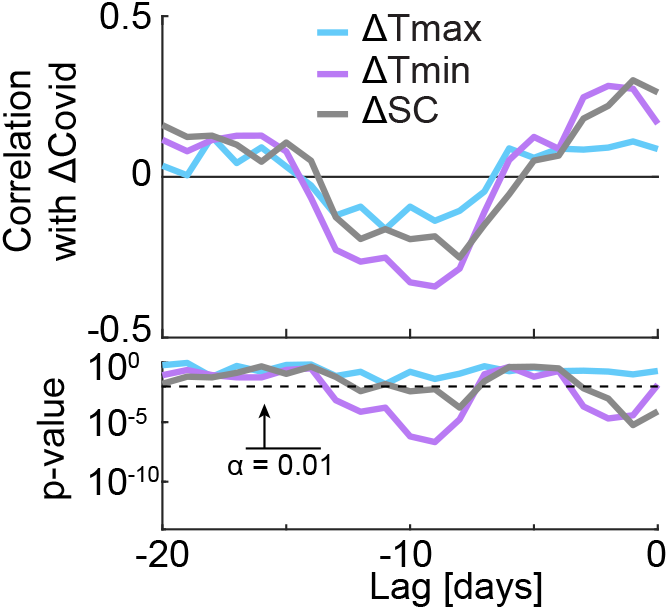
As Figure 1B but for the remaining meteorological variables, not shown there to allow for visual clarity.

**Figure S5.**
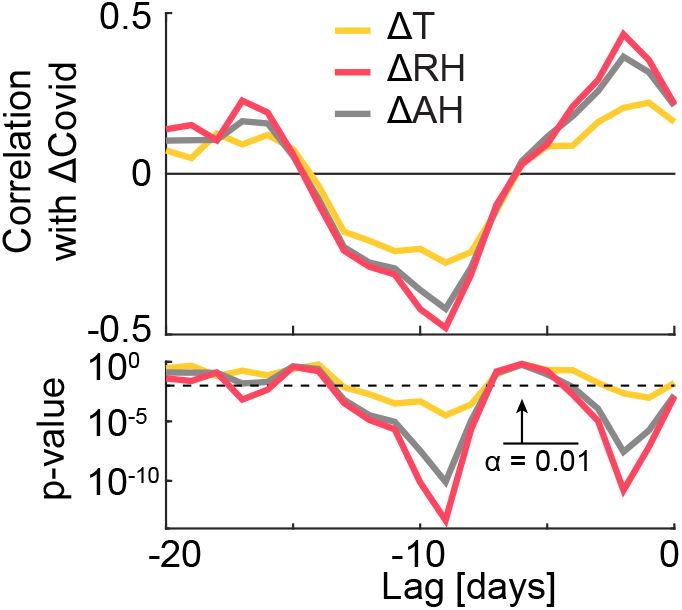
As Figure 1B but showing ΔT (orange), ΔRH (red) and the estimation of ΔAH obtained with Equation 1 (grey).

